# Genome-wide association study identifies risk variants for sporadic Creutzfeldt-Jakob disease in *STX6* and *GAL3ST1*

**DOI:** 10.1101/2020.04.06.20055376

**Authors:** Emma Jones, Holger Hummerich, Emmanuelle Viré, James Uphill, Athanasios Dimitriadis, Helen Speedy, Tracy Campbell, Penny Norsworthy, Liam Quinn, Jerome Whitfield, Jacqueline Linehan, Zane Jaunmuktane, Sebastian Brandner, Parmjit Jat, Akin Nihat, Tze How Mok, Parvin Ahmed, Steven Collins, Christiane Stehmann, Shannon Sarros, Gabor Kovacs, Michael Geschwind, Aili Golubjatnikov, Karl Fronztek, Herbert Budka, Adriano Aguzzi, Hata Karamujić-Čomić, Sven van der Lee, Carla A Ibrahim-Verbaas, Cornelia Van Duijn, Beata Sikorska, Ewa Golanska, Pawel Liberski, Miguel Calero, Olga Calero, Pascual Sanchez Juan, Antonio Salas, Federico Martinón-Torres, Elodie Bouaziz-Amar, Stephane Haik, Jean-Louis Laplanche, Jean-Phillipe Brandel, Phillipe Amouyel, Jean-Charles Lambert, Piero Parchi, Anna Bartoletti-Stella, Sabina Capellari, Anna Poleggi, Anna Ladogana, Maurizio Pocchiari, Serena Aneli, Giuseppe Matullo, Richard Knight, Saima Zafar, Inga Zerr, Stephanie Booth, Michael B Coulthart, Gerard H Jansen, Katie Glisic, Janis Blevins, Pierluigi Gambetti, Jiri Safar, Brian Appleby, John Collinge, Simon Mead

**Affiliations:** MRC Prion Unit at UCL, Institute of Prion Diseases, 33 Cleveland St, London, W1W 7FF; National Prion Clinic, University College London Hospitals NHS Foundation Trust, London; Division of Neuropathology, University College London Hospitals NHS Foundation Trust, and Department of Clinical and Movement Neurosciences and Queen Square Brain Bank for Neurological Disorders, UCL Queen Square Institute of Neurology, London, UK; Division of Neuropathology, University College London Hospitals NHS Foundation Trust, and Department of Neurodegenerative disease, UCL Queen Square Institute of Neurology, Queen Square, London WC1N 3BG; Australian National Creutzfeldt-Jakob Disease Registry, Florey Institute of Neuroscience and Mental Health, The University of Melbourne, Victoria, 3010, Australia; Institute of Neurology, Medical University of Vienna, Vienna, Austria; Department of Laboratory Medicine and Pathobiology and Tanz Centre for Research in Neurodegenerative Disease, University of Toronto, and Laboratory Medicine Program & Krembil Brain Institute, University Health Network, Toronto, Ontario Canada; UCSF Memory and Aging Center, Department of Neurology, University of California, San Francisco; Institute of Neuropathology, University of Zurich, Zurich, Switzerland; Department of Epidemiology, Erasmus Medical Centre, Rotterdam, the Netherlands; Nuffield Department of Population Health, University of Oxford, UK; Laboratory of Electron Microscopy and Neuropathology, Department of Molecular Pathology and Neuropathology, Medical University of Lodz, Lodz, Poland; Chronic Disease Programme (UFIEC-CROSADIS) and Network Center for Biomedical Research in Neurodegenerative Diseases (CIBERNED), Instituto de Salud Carlos III, Madrid, Spain and Alzheimer Disease Research Unit, CIEN Foundation, Queen Sofia Foundation Alzheimer Center, Chronic Disease Programme Carlos III Institute of Health and Network Center for Biomedical Research in Neurodegenerative Diseases (CIBERNED), Madrid, Spain; Neurology Service, University Hospital Marqués de Valdecilla (University of Cantabria, CIBERNED and IDIVAL), Santander, Spain; Unidade de Xenética, Instituto de Ciencias Forenses (INCIFOR), Facultade de Medicina, Universidade de Santiago de Compostela, and GenPoB Research Group, Instituto de Investigaciones Sanitarias (IDIS), Hospital Clínico Universitario de Santiago (SERGAS), Galicia, Spain; Translational Pediatrics and Infectious Diseases, Department of Pediatrics, Hospital Clínico Universitario de Santiago de Compostela, Galicia, Spain; Department of Biochemistry and Molecular Biology, Lariboisière Hospital, AP-HP, University of Paris, France; Sorbonne Université, INSERM, CNRS UMR 7225, Institut du Cerveau et de la Moelle épinière, ICM, Paris, France; Cellule nationale de référence des maladies de Creutzfeldt-Jakob, APHP, University Hospital Pitié-Salpêtrière, Paris, France; Univ. Lille, Inserm, CHU Lille, Institut Pasteur de Lille, U1167-RID-AGE, Labex DISTALZ, Lille, France; IRCCS, Istituto delle Scienze Neurologiche di Bologna, Bologna, Italy; Department of Biomedical and Neuromotor Sciences, University of Bologna, Bologna, Italy; Department of Experimental, Diagnostic and Specialty Medicine, University of Bologna, Bologna, Italy; Department of Neuroscience, Istituto Superiore di Sanità, Rome, Italy; Department of Medical Sciences, Università degli studi di Torino, Via Verdi, Torino, Italy; National CJD Research and Surveillance Unit, Edinburgh, UK; Department of Neurology, Clinical Dementia Center and National Reference Center for CJD Surveillance, University Medical School; German Center for Neurodegenerative Diseases (DZNE), Göttingen, Germany; Prion Disease Program, Public Health Agency of Canada, Winnipeg, Canada; Canadian Creutzfeldt-Jakob Disease Surveillance System, Public Health Agency of Canada, Ottawa, Canada; Department of Pathology and Laboratory Medicine, University of Ottawa, Ottawa, Canada; Departments of Pathology and Neurology, Case Western Reserve University, Cleveland, OH, USA; National Prion Disease Pathology Surveillance Center, Case Western Reserve University, Cleveland, OH, USA

**Keywords:** prion, GWAS, CJD, STX6, GAL3ST1

## Abstract

Mammalian prions are lethal pathogens composed of fibrillar assemblies of misfolded prion protein. Human prion diseases are rare and usually rapidly fatal neurodegenerative disorders, the most common being sporadic Creutzfeldt-Jakob disease (sCJD). Variants in the gene that encodes prion protein (*PRNP*) are strong risk factors for sCJD, but although the condition has heritability similar to other neurodegenerative disorders, no other risk loci have yet been confirmed. By genome-wide association in European ancestry populations, we found three replicated loci (cases n=5208, within *PRNP, STX6*, and *GAL3ST1*) and two further unreplicated loci were significant in gene-wide tests (within *PDIA4, BMERB1*). Exome sequencing in 407 sCJD cases, conditional and transcription analyses suggest that associations at *PRNP* and *GAL3ST1* are likely to be caused by common variants that alter the protein sequence, whereas risk variants in *STX6* and *PDIA4* associate with increased expression of the major transcripts in disease-relevant brain regions. Alteration of *STX6* expression does not modify prion propagation in a neuroblastoma cell model of mouse prion infection. We went on to analyse the proteins histologically in diseased tissue and examine the effects of risk variants on clinical phenotypes using deep longitudinal clinical cohort data. Risk SNPs in *STX6*, a protein involved in the intracellular trafficking of proteins and vesicles, are shared with progressive supranuclear palsy, a neurodegenerative disease associated with the misfolded protein tau. We present the first evidence of statistically robust associations in sporadic human prion disease that implicate intracellular trafficking and sphingolipid metabolism.

## Introduction

Prion diseases are fatal neurodegenerative conditions of humans and animals caused by the propagation of prions, atypical infectious agents comprised solely or predominantly of host prion protein^1^. Prions are thought to propagate through a process of binding to normal prion protein, induction of conformational change by templating and fission of the polymeric assembly. Prion diseases can be acquired by exposure to prions in the diet, or through medical or surgical procedures, which may result in public health crises. The cattle prion disease, bovine spongiform encephalopathy (BSE), which transmitted to mostly young British and other European adults as variant Creutzfeldt-Jakob disease (vCJD)^2^, led to enhanced clinical surveillance for all prion diseases worldwide. Inherited prion disease, caused only by mutations of the prion protein (*PRNP*) gene, causes approximately 10-15% of the annual incidence in most countries^3^. By far the most common type, however, is sporadic CJD (sCJD), a rapidly progressive dementia with a lifetime risk of approximately 1 in 5000, which occurs predominantly in older adults^4^. Other than age and variation at *PRNP*, no risk factors for sCJD are known, leaving only speculative explanations for why prions might spontaneously develop in an older person.

Polymorphisms of *PRNP* that alter the amino-acids at codons 127, 129 and 219 are known to be strong genetic risk factors for, or modifiers of, the phenotypes of prion diseases^3^. Sibling or familial concurrence of sCJD has been reported, but not to the extent that chance concurrence can be eliminated as an explanation. There are also no estimates of the heritability of sCJD based on family studies^5^. Animal studies have identified risk factors for acquired prion diseases in and close to *Prnp*, and evidence for susceptibility loci on other chromosomes, although studies to identify the causal genes have been challenging^3^. This study follows on from previous genome-wide association studies (GWAS) in human prion diseases, which have not been powerful enough to discover non-*PRNP* risk factors^6-8^. Many neurodegenerative diseases are thought to share fundamental mechanisms with prion diseases invoking template-based protein misfolding, and spreading of pathology associated with abnormally aggregated proteins in diseased brain tissue. If shared mechanisms exist this might implicate joint genetic risk factors.

Here we conducted a GWAS in sCJD, based on samples largely derived from clinical surveillance in countries with populations of predominantly European ancestries. We went on to do correlations with clinical phenotypes, transcriptional, protein and cell model-based analyses with the aim of supporting the specific causal genes at risk loci and allowing propositions of molecular mechanisms. We identified new risk factors for sCJD including variants which appear to have pleiotropic effects in neurodegenerative diseases.

## Results

In the discovery phase we generated genome-wide genotype data from 4110 patients with probable or definite sCJD, from nine countries of predominantly European ancestries and we compared them with 13,569 control samples from seven countries (see Methods, Supplementary Tables 1 and 2 for array types, age, sex, and clinical phenotype data). Imputation using the Michigan server resulted in 6,314,477 high quality autosomal SNPs after QC (see methods) which were used for downstream association tests in SNPTEST with 10 population covariates. Genomic inflation (λ) was 1.026 (see QQ plots with and without GWAS significant loci, Supplementary Figure 1), indicating no significant and systemic bias related to population ancestry or platforms, so no further correction was done; the threshold for genome-wide significance was P<5×10^−8^. Both SumHer (h^2^_SNP_ =0.26) and GCTA (h^2^ _SNP_=0.25) estimated similar SNP heritability to common neurodegenerative diseases, in keeping with only very rare reports of familial concurrence of sCJD^5,9,10^.

Further to the known association at *PRNP* on chromosome 20p13, two loci achieved genome-wide significance mapping to 1q25.3 (*STX6*) and 22q12.2 (*GAL3ST1*) (see Manhattan plot, Figure 1; Table 1; Supplementary Figures 2, 3 and 4 for regional association plots). Gene based testing using MAGMA and VEGAS additionally identified *PDIA4* (P=0.040 (VEGAS2) and no significant result for MAGMA)) and *BMERB1* (P=0.0014 (VEGAS2) and no significant result for MAGMA) as showing gene-wide significance (Supplementary Figures 5 and 6). No gene-sets were significant. A SNP in intron 1 of the *BMERB1* gene achieved borderline significance (rs6498552, P=5.75 × 10^−8^, Supplementary Figure 6). A lead SNP from the three genome-wide significant loci and a lead SNP from *PDIA4* and rs6498552 in *BMERB1* gene were taken into a replication phase, whilst we acknowledge that data from multiple SNPs at a locus are needed to directly replicate gene-based test results.

**Table 1.**
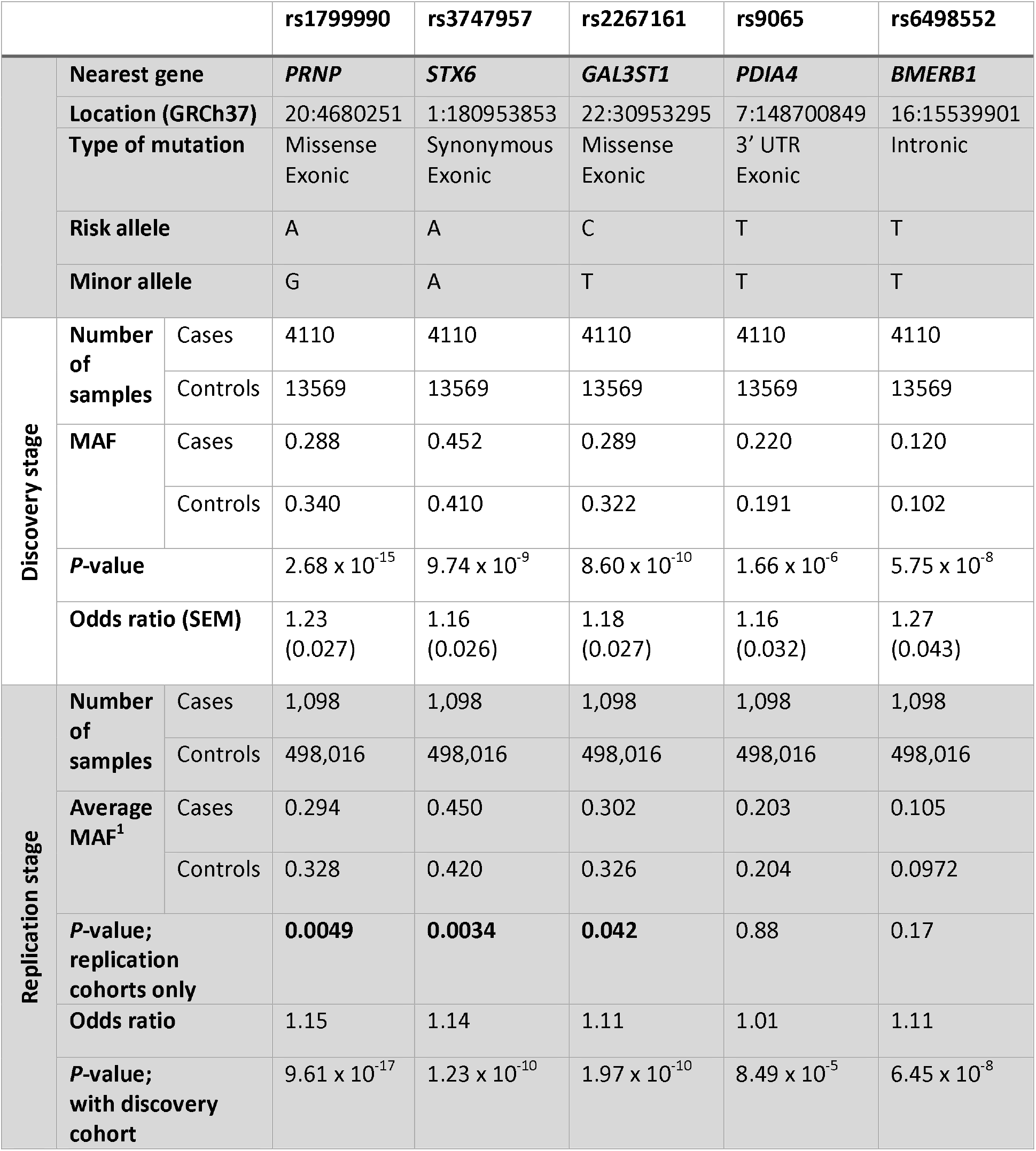
Main results of discovery and replication stages. *PRNP, STX6* and *GAL3ST1* SNPs were successfully replicated in an independent cohort (P < 0.05) with a similar effect as the discovery phase. ^1^Replication minor allele frequencies (MAF) were estimated as a weighted mean of each cohort based on case sample number.

**Figure 1.**
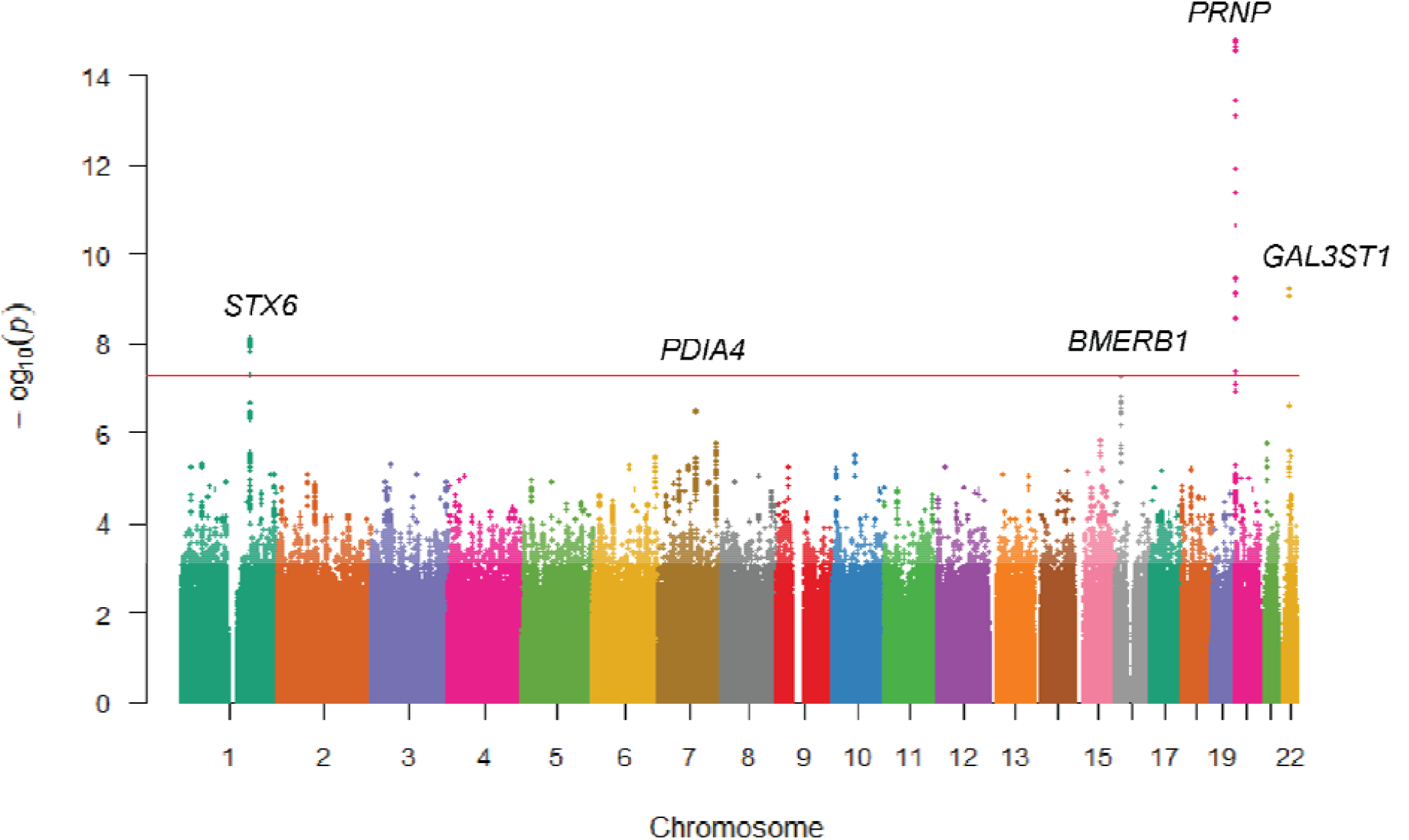
Manhattan plot. Illustrating three genome-wide significant loci conferring risk of sCJD (in *PRNP, STX6* and *GAL3ST1*) and two genes significant only with gene-wide tests (*PDIA4, BMERB1*).

In the replication phase we generated genotype data using minor groove binding probes from 1098 patients with probable or definite CJD, from seven countries of predominantly European ancestries and compared with genotypes from 498,016 control samples from the same seven countries (see Methods, Supplementary Tables 1 and 2). Association testing provided replication evidence for *PRNP* (rs1799990, heterozygous genotype and to a lesser extent the minor allele is protective), *STX6* (rs3747957, minor allele conferred risk), and *GAL3ST1* (rs2267161, minor allele was protective), but not individual lead SNPs in *PDIA4*, or *BMERB1* (Table 1). We went on to attempt to extend evidence of an association in related prion diseases including vCJD, acquired from exposure to BSE; iatrogenic CJD caused by exposure to cadaveric pituitary-derived human growth hormone; or kuru (and resistance to kuru), a former epidemic orally transmitted prion disease of people who lived in the Eastern Highlands Province of Papua New Guinea (see Methods). We found no evidence for association of rs3747957 in *STX6*, or rs2267161 in *GAL3ST1* with these phenotypes (P>0.05), implying that these loci confer risk specific to the sporadic form of human prion disease, although sample sizes for these conditions are much smaller than for sCJD and all tests were underpowered^6,8^.

Sporadic CJD is known to comprise a range of different clinical and pathological phenotypes, broadly correlating with prion molecular strain types, the latter including categorisation by different proportions of three glycoforms and the apparent molecular weight of abnormal PrP by western blotting^11^. For 10 years at the National Prion Clinic London we have conducted longitudinal observational cohort studies of CJD, involving systematic clinical assessments of patients resulting in deep phenotype data^12,13^. We tested rs1799990, rs3747957 and rs2267161 for association with age at clinical onset, clinical duration, the slope of decline in a functional measure of disease severity, along with 27 other phenotypic variables (Supplementary Table 3). Only rs1799990 in *PRNP* showed, as expected, associations with several clinical and biomarker traits (10/30 tested hypotheses). There was no evidence for epistasis between discovered loci and genotypes at rs1799990, which encodes different primary sequences of PrP and is known to be a major determinant of clinical phenotype and categories.

Associations in a genomic region might not implicate effects through the nearest gene. We therefore used tools to investigate the potential mechanisms underlying associations at the genomic regions of *PRNP, STX6* and *GAL3ST1*. Initially, we used CAVIAR, a method of fine mapping of the association signal at a locus through jointly modelling association statistics for all variants at a locus, then estimating a conditional posterior probability of causality whilst allowing for multiple plausibly functional SNPs^14^. For *PRNP*, SNPs tagging rs1799990 were predominantly identified, as expected, however a cluster of SNPs 5’ (lead SNP rs12624635, not an eQTL) to these with low LD to rs1799990 were also putatively causal suggesting a potential additional signal at this locus (Figure 2a). Previous studies have reported that variants at the *PRNP* locus may confer an increase in sCJD risk, independent to the effect at rs1799990^15-18^. However, in a conditional analysis, adjusting for heterozygosity at rs1799990, we found no substantive evidence of an independent association signal (Supplementary Figure 7 and 8); lead SNP rs12624635, P=0.03 (heterozygous conditional model).

**Figure 2.**
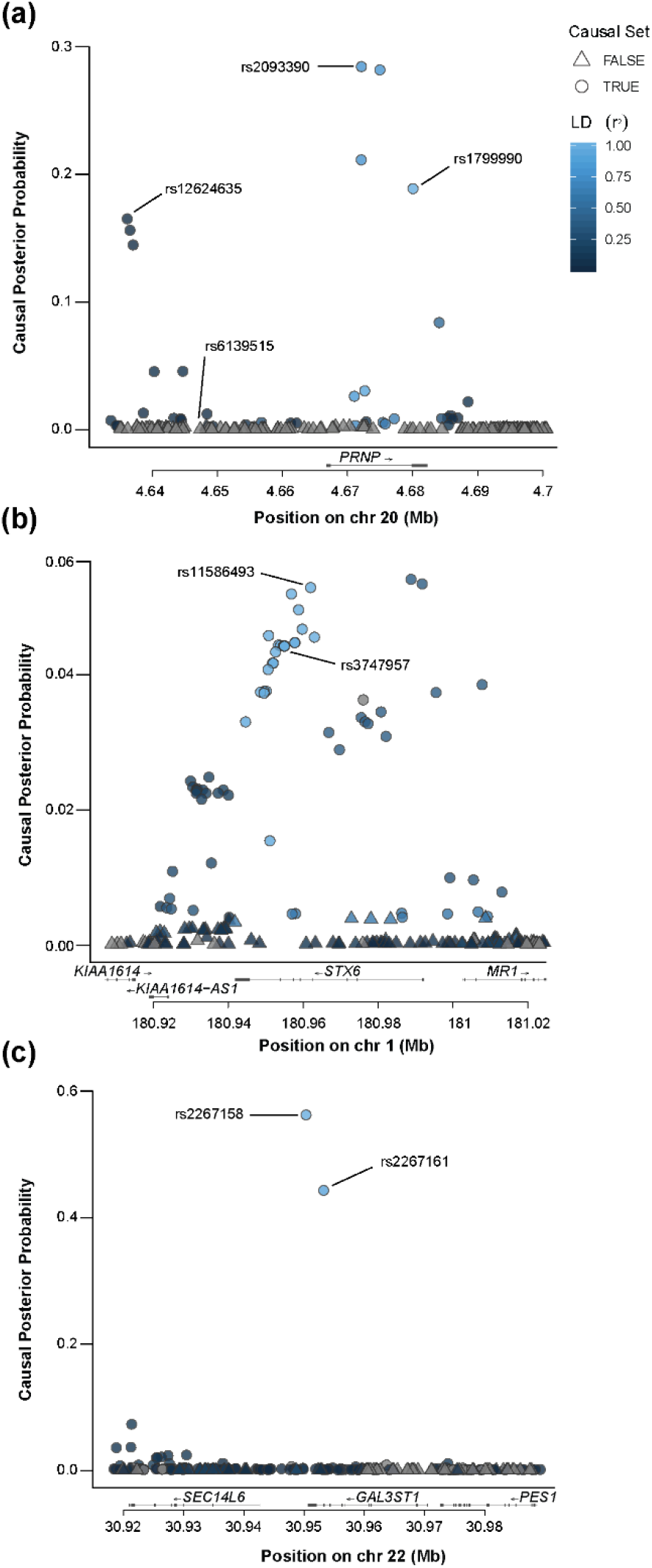
Statistical fine-mapping using CAVIAR identifies set of putative causal variants at each GWAS significant locus, suggesting potential additional signal in *PRNP* upstream of codon 129. CAVIAR utilizes summary statistics and LD structure to predict the probability of each variant being causal, producing a ‘causal set’ with 95% probability of containing the causal SNP, whilst allowing for the possibility of multiple causal SNPs at each locus. Each locus was defined as 100 variants upstream and downstream of the top SNP. Plots show causal posterior probability of each variant at (**a**) *PRNP*, (**b**) *STX6* and (**c**) *GAL3ST1* coloured by LD (1000G; EUR) with the top SNP. Circles indicate variants within the 95% causal set.

The region of high LD surrounding rs3747957 in *STX6* results in a large causal set, making identification of a single causal variant more complex (Figure 2b). Subsequently, using eCAVIAR and GTEx^19^ and other eQTL databases, we identified a strong correlation between sCJD risk and increased expression of *STX6* mRNA in multiple brain regions, particularly in the caudate and putamen nuclei of the brain (Putamen STX6 rs3747957 P=2.3 × 10^−13^, GTEx, Figure 3), key regions implicated in sCJD and the most frequently abnormal brain regions at diagnostic MRI brain imaging^20^. Correlations between lead SNPs in *STX6* rs11586493 and rs3747957 and other genes at the locus or within other tissues were absent or less strong (Figure 3, Supplementary Table 4). These analyses suggest that increased expression of *STX6* in brain regions confers an increased risk of sCJD. eQTLs for nearby gene *KIAA1614* in the tibial nerve (but not brain tissues) were also co-localised to a less extent with the GWAS signal; this encodes an uncharacterised protein with little known about its cellular function, however a disease-related mechanism cannot be ruled out. Analysis with PAINTOR, a tool that integrates functional genomic annotation with association statistics, identified three SNPs (rs12754041, rs10797664 and rs6425657, each in strong LD with lead SNPs at the locus) as having a high posterior probability of being causal in four annotation groups (RoadMap_Assayed_NarrowPeak; Maurano_Science2012_DHS; RoadMap_Enhancers; Roadmap_ChromeHMM_15state) (see Methods)^21^.

**Figure 3.**
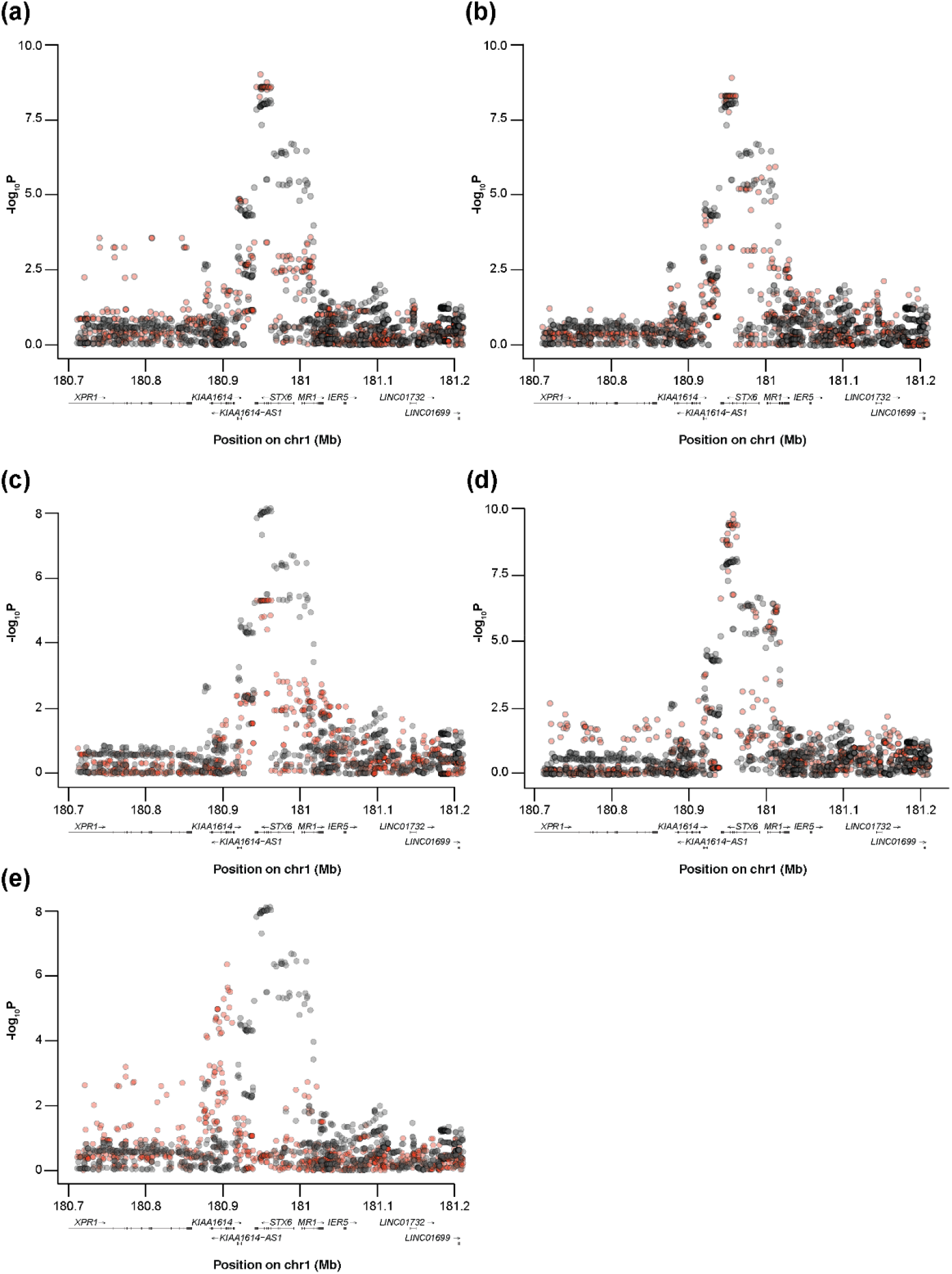
GWAS results for sCJD at *STX6* co-localise with eQTLs for *STX6* expression in the caudate, putamen and hypothalamus, and *KIAA1614* expression in the tibial nerve. Plot of –log_10_ of P-values from the GWAS association analysis at the *STX6* locus (black) and the eQTL association analysis from the GTEx dataset (v7) (red) for: (**a**) *STX6* expression in the caudate, (**b**) *STX6* expression in the putamen, (**c**) *STX6* expression in the hypothalamus, (**d**) *KIAA1614* expression in the tibial nerve, (**e**) *KIAA1614* expression in the tibial artery. Peaks correspond to the CLPP in the eCAVIAR analysis with a higher degree of colocalisation with increasing CLPP (see Supplementary Table 4).

As the GWAS signal is associated with only two SNPs at *GAL3ST1* (in strong LD with each other but low LD with all other surrounding variants) these SNPs predominantly define the causal set as expected, however, these are statistically indistinguishable from each other (Figure 2c). One of these SNPs, rs2267161, is a non-synonymous variant of *GAL3ST1* p.V29M. In GTEx, neither SNP correlates with expression of genes at the locus in brain tissues. Close to p.V29M resides p.V34M (rs55674628, allele frequency=0.02; LD with rs2267161, r^2^=0.01, D’=1.00, discovery P=0.18), these being the only common non-synonymous variants in European ancestries populations. These polymorphisms form three common haplotypes, rs2267161-C/rs55674628-C (CC), CT and TC with frequencies in the combined case-control dataset of 0.667, 0.018 and 0.315, respectively. In a haplotype-based test, no evidence of an association driven by the rs55674628-T allele was observed (Supplementary Table 5). No additional rare variants of *GAL3ST1* or *STX6* were observed in the exomes of 407 sCJD cases^22^.

Expression of *STX6, GAL3ST1, PDIA4* and *BMERB1* mRNA was slightly reduced in bulk analysis of post-mortem cerebellar brain tissue from sCJD cases but only to a similar extent as genes suggested to be good comparators (Supplementary Figure 9)^23^. By immunohistology in the frontal cortex (19 sCJD, 15 control cases, see Methods), syntaxin-6 staining appears restricted to neurons of different sizes, whilst other cell types, probably astrocytes or oligodendrocytes, were less consistently stained. In the cerebellum, the staining is seen in Purkinje cells, in large neurons of the dentate nucleus, and there is also a fine, granular staining within the molecular layer (Supplementary Figure 10a). In all neuron populations of cerebellum and forebrain, the staining pattern is fine granular, and is located in the cytoplasm, but does not extend into the processes. The staining pattern is compatible with the predicted target, the Golgi apparatus. The pattern was indistinguishable between CJD cases and controls. See Supplementary Figure 10b and legend for Protein Disulphide Isomerase Family A Member 4 immunohistology.

As we had a clear hypothesis based on GTEx data, that increased expression of *Stx6* in deep brain nuclei increases risk of prion disease, we wished to test the suggestion that risk might be conferred through syntaxin-6 facilitating prion propagation in mammalian neuronal cells. We therefore used RNA interference to stably knockdown *Stx6* expression in prion-susceptible mouse neuroblastoma-derived cells (N2aPK1/2)^24^ and measured impact on prion propagation in the Automated Scrapie Cell Assay^24,25^. Cells were exposed to Rocky Mountain Laboratory (RML) prions and resultant spot count was compared to *Prnp* knockdown cells previously demonstrated to inhibit prion propagation in this assay (Figure 4)^26^, as well as control cells transfected with a scrambled shRNA sequence. Despite similar efficiency of knockdown in these cell lines a consistent effect on prion propagation was not established suggesting, unlike *Prnp*, reduced expression of *Stx6* does not have a consistent effect on the ability of these cells to propagate RML prions.

**Figure 4:**
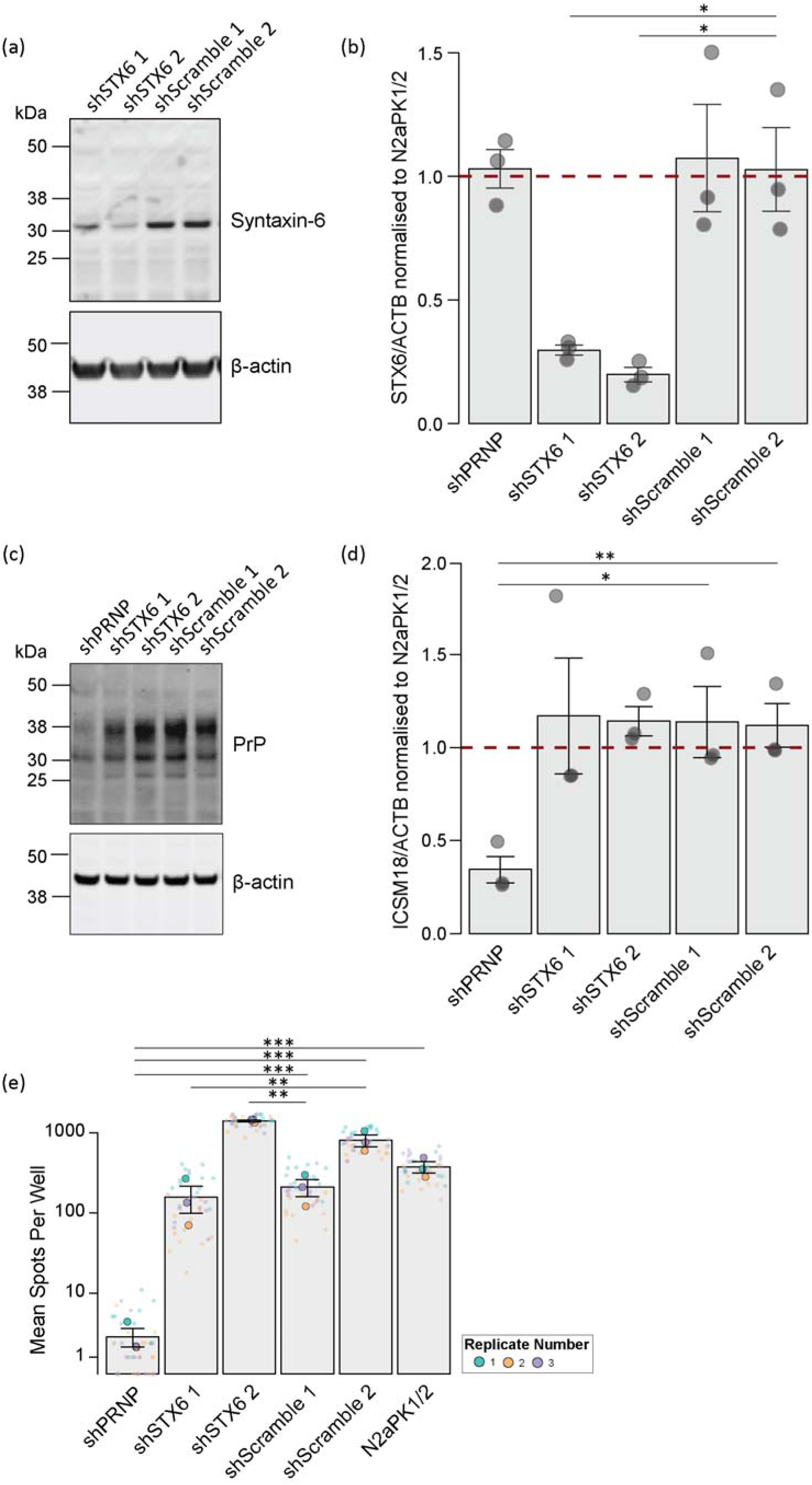
Knockdown of *Stx6* expression in N2aPK1/2 cells by >70% does not have a consistent effect on prion propagation in the scrapie cell assay (SCA). N2aPK1/2 cells were transfected with pRetroSuper vectors containing *Stx6* (‘shSTX6 1’, ‘shSTX6 2’) or *Prnp* (‘shPRNP’) targeting shRNAs to generate knockdown cell lines or a scrambled non-silencing (‘shScramble 1’, ‘shScramble 2’) shRNA sequence for controls. Samples were taken prior to plating for SCA for immunoblot (n = 3) and expression normalised to untransfected N2aPK1/2 (indicated by dashed line). (**a-b**) Knockdown of syntaxin-6 protein levels determined by (**a**) immunoblot with anti-syntaxin-6 antibody with (**b**) band intensity measured relative to β-actin loading control (Student’s t-test). (**c-d**) Knockdown of PrP^C^ protein determined by (**c**) immunoblot with anti-PrP antibody ICSM18 with (d) band intensity measured relative to β-actin loading control (Student’s t-test). (e) Average spot count of infected cell number post-4^th^ split in SCA following infection with RML at 3 × 10^−6^ dilution (one-way ANOVA with Tukey’s post-hoc test on log-transformed data). Statistical associations of knockdown lines relative to controls indicated; other results omitted for clarity. All error bars show mean ± SEM; *P < 0.05, ** P < 0.01, *** P < 0.001.

## Discussion

We report the first GWAS in a human prion disease powered to detect alleles with the modest effects typical of complex diseases. Further to the known effects at *PRNP* codon 129, we report two independently replicated loci, and evidence to support the conclusion that risk variants modify the primary sequence of the encoded protein (*GAL3ST1*) or increase expression level in brain tissues (*STX6*). A multitude of potential binding partners for PrP and mechanisms for the modification of prion infection have been proposed. GWAS discoveries have special value because risk variants are implicitly causal in the human disease^27^. Therapeutic targets underpinned by genetic evidence have better chances of successful drug development, which should encourage research efforts to further understanding the mechanisms that underpin these signals^27^.

Risk variants in sCJD might act at different stages, for example, increasing the chance of the spontaneous generation of prions, or reducing their clearance, enabling prion propagation throughout brain tissue, or the downstream toxic effects of prion propagation on brain cells. Whilst it is too early to draw confident conclusions in this respect as power was low, we did not find any evidence of a role for risk variants in the modification of clinical or pathological disease phenotypes, or in modified expression of risk genes at the end stage of the disease process. Alteration of the expression level of *STX6* in a cellular model of prion infection did not modify the susceptibility of these cells to infection or the accumulation of abnormal forms of PrP. Our initial functional data, therefore, point to a role early in the disease process, perhaps by altering the risk of spontaneous prion formation in the human brain, however clearly, studies in other neuronal models and animals are warranted.

*STX6* encodes syntaxin-6, an 11 exon, 255 amino-acid protein that localises to the trans-Golgi network, recycling and early endosomes. Syntaxin-6 is thought to form part of the t-SNARE complex involved in the decision of a target membrane to accept the fusion of a vesicle^28^. The intracellular location of abnormal PrP in prion-infected cells involves the plasma membrane where conversion is primarily thought to occur^26^, as well as early and recycling endosomes, late endosomes and the perinuclear region^29^. Other studies implicate the endocytic-recycling compartments (ERCs) and/or multivesicular bodies (MVBs) for the sites of generation of prions, and dysregulation of trafficking genes by sCJD^30,31^. Intracellular trafficking has also been implicated in the degradation of prions^32^. The modification of trafficking of normal and/or abnormal PrP by syntaxin-6 might be a focus for future investigation.

There has been considerable recent discussion about the extent to which neurodegenerative diseases associated with the accumulation of misfolded forms of normal proteins or peptides are “prion-like” in their pathogenesis^33^. This concept provokes the suggestion that prion and prion-like disorders might share genetic risk factors. Progressive supranuclear palsy (PSP) is an uncommon neurodegenerative movement disorder, associated with the accumulation of abnormal forms of microtubule-associated protein tau (tau) with four repeats^34,35^. Variants in *STX6* are shared risk SNPs with a similar direction of effect in sCJD and PSP^36^. Pleiotropic effects at this locus between prion diseases and a tauopathy lend support to the concept of prion-like disorders and raise the prospect for genetically inspired interventions across multiple neurodegenerative disorders.

*GAL3ST1* encodes galactose-3-O-sulfotransferase 1, a 423 amino acid protein that localises to the Golgi network in oligodendrocytes, the sole enzyme responsible for the sulfation of membrane sphingolipids to form sulfatides, a major brain lipid and component of the myelin sheath^37^. Degradation of sulfatides is catalysed by *ARSA* in the lysosome; recessive defects in this enzyme cause metachromatic leukodystrophy, a lysosomal storage disorder associated with profound central and peripheral demyelination^38^. Knockout of *GAL3ST1* in mice results in a neurological phenotype associated with abnormal myelin maintenance with age, and histological abnormalities at the paranodal junctions^39^. Sphingolipid metabolism and myelin maintenance have both been previously implicated in PrP and prion diseases^40,41^. Multiple genes in the sphingolipid metabolic pathways are dysregulated early in the pathogenesis of mouse prion diseases, a finding consistent between inbred strains and prion strains^42^. Knockout of PrP in mouse, or naturally in goats, results in a demyelinating neuropathy, which in goats is associated with abnormal sphingolipid metabolism^43-45^.

*PDIA4* and *BMERB1* loci identified in the discovery phase by gene-wide analysis, were not replicated by single SNP tests, however, the replication sample was necessarily limited by the rarity of the disease, the lead SNPs had a lower allele frequency than at other risk loci, and further attempts are justified as gene-based test results are driven by multiple SNPs at each locus. We note that *PDIA4* is associated with the unfolded protein response, implicated in prion disease pathogenesis, and the gene family is increased in expression by the disease, as well as some evidence for a direct interaction with PrP^46^. Variants in *PDIA4* most strongly associated with disease, are also eQTLs for brain expression of the gene (GTEx). Little is known about *BMERB1* (an uncharacterised protein showing bivalent Mical/EHBP Rab binding) but by homology may have a role in intracellular trafficking^47^.

In summary, we present the first evidence of statistically robust associations in sporadic human prion disease that implicate intracellular trafficking and sphingolipid metabolism.

## Materials and methods

### Human Samples Selection

Samples from patients with prion diseases were provided by specialist or national surveillance centres in countries with populations of predominantly European ancestries. A diagnosis of probable or definite sporadic CJD was required according to the contemporary widely accepted criteria (for most recent example see https://www.cjd.ed.ac.uk/sites/default/files/NCJDRSU%20surveillance%20protocol-january%202017.pdf). These criteria have evolved over time, principally to include the recognition of the importance of MRI brain imaging, and the cerebrospinal fluid Real-Time Quaking Induced Conversion Assay (RT-QuIC) in diagnosis. As there was no restriction on the calendar date of diagnosis many patients were diagnosed using previous versions of diagnostic criteria with similarly high levels of specificity. Circumstances particular to each contributing site are detailed below, the activity and methods of surveillance for CJD vary between countries. For average age of clinical onset, % female, clinical duration, and proportion with a pathological diagnosis, see Supplementary Table 2. Ethical approval for research studies was provided by London – Harrow Research Ethics Committee.

### London, UK

Samples from patients with probable or definite sCJD were acquired through the activities of the National Prion Clinic, London and National CJD Research and Surveillance Unit, Edinburgh. Both Units coordinate their work closely and samples for research are shared according to a National Agreement, 2004. The laboratory genetic studies were approved by the London Harrow Research Ethics Committee. The National Prion Monitoring Cohort Study was approved by the Scotland A Research Ethics Committee. Samples from patients with probable or definite variant CJD were acquired from both Units and were used in follow up analyses of loci identified by sCJD GWAS. 119 samples from patients with probable or definite variant CJD, and 38 with probable or definite iatrogenic CJD were acquired through the activities of the National Prion Clinic, London and National CJD Research and Surveillance Unit, Edinburgh. These were processed for association testing in as per sCJD (see Quality control and imputation) and compared with UK controls.

### Austria

The Austrian CJD Surveillance is supported by the Ministry of Health of Austria (OERPE). Patients with sporadic CJD were referred by clinical neurologists from various clinics and finally ascertained by the National CJD Surveillance Center. Classification of cases was according to WHO criteria updated to include MRI features^48^ and all cases underwent neuropathological examination and immunostaining for disease-associated PrP. Research studies were approved by the Ethics Committee of the Medical University of Vienna (396/2011, updated every year).

### Australia

The Australian National Creutzfeldt-Jakob Disease Registry (ANCJDR) is funded by the Commonwealth Department of Health with surveillance methods to ascertain patients with sporadic CJD previously detailed^49^. Classification of cases was according to WHO criteria updated to include MRI features^48^, with permission for inclusion in research studies as described^50^.

### Canada

The Canadian National CJD surveillance System (CJDSS) is funded by the Public Health Agency of Canada (PHAC) to perform laboratory reference services for neuropathology and molecular genetic investigation of suspected prion disease. The WHO internationally accepted model for comprehensive surveillance of human prion diseases is used in the classification of cases. Samples from patients with pathologically confirmed CJD with consent for inclusion in research studies were acquired. Ethical approval for genetic studies were approved by the Health Canada and PHAC Research Ethics Board.

### France

These samples were duplicated from a previous GWAS in sCJD using a different genome-wide array. See^7^ for details.

### Netherlands

The Dutch National Prion Disease Registry is located at the Erasmus MC, University Medical Center, Rotterdam, The Netherlands. In the Netherlands, it is obligatory by law to report patients suspected of CJD or other prion disease to the National Prion Disease Registry. The National Prion Disease Registry collects detailed information from all patients with regards to the clinical disease course, medical history, and results from diagnostic information in order to estimate the probability of the prion disease and the type of the prion disease. Besides the obligatory part of the National Prion Disease Registry, the patients and families are asked for consent to participate in a genetic-epidemiologic research study, which is performed by the physician from the National Prion Disease Registry. This research study consists of an extended questionnaire, blood withdrawal from patients and relatives, and collection of data from the medical records. All patients and relatives participating in this study provided written informed consent to participate and to obtain information from their treating physicians. The patients included in this study were probable sCJD cases or definite sCJD cases based on the diagnostic criteria from the University of Edinburgh.

### Germany

In Germany, patients were assessed and diagnosed by members of the National Reference Center for TSE, University Medical Center Goettingen in the framework of a prospective epidemiological study including genetic research. The study has been approved by the local ethics committee. The patients fulfilled clinical WHO criteria for probable sporadic CJD or were confirmed as definite CJD by neuropathological examination. New German samples were added to those previously studied definite CJD cases from the Center for Neuropathology and Prion Research, Munich^51^.

### Italy

Patients clinically suspected of having CJD were referred to the surveillance system and diagnosed at last follow-up as probable or definite sporadic CJD according to the current European diagnostic criteria. Informed written consent from patients or next of kin was obtained for assessing the presence of *PRNP* mutation to exclude genetic forms of prion disease.

### Poland

Patients with sporadic CJD were referred by clinical neurologists from various clinics and finally assessed by EG, BS, PPL according to the WHO criteria for probably sporadic CJD, or at post-mortem examination by BS, Department of Molecular Pathology and Neuropathology. Approval for genetic research studies was given by the local research ethics committee.

### Spain

All samples for sCJD cases analysed were obtained from patients of Spanish origin with suspected prion diseases, submitted for diagnostic purposes under the guidelines of the Spanish National Referral and Surveillance system. The study was approved by the Bioethics and Animal Welfare Committee from the Instituto de Salud Carlos III, Madrid, Spain. For this study, all samples were coded and personal information dissociated from the test results, according to local legislation at the time of analysis. All the data were analysed anonymously, and all clinical investigations have been conducted according to the principles expressed in the Declaration of Helsinki.

### Switzerland

Irreversibly anonymized samples collected between December 1996 and May 2012 were obtained from the tissue bank of the Swiss National Reference Center for Prion Diseases (University of Zurich, Zurich, Switzerland). Suspected cases underwent western blot and/or immunohistochemical analyses, therefore, all patients fulfilled the WHO criteria for definite sporadic CJD. The study was approved by the local research ethics committee.

### National Prion Disease Pathology Surveillance Centre and UCSF, USA

The National Prion Disease Pathology Surveillance Centre (NPDPSC) is funded by the Centres for Disease Control and Prevention (CDC) to perform neuropathologic surveillance of cases of suspected prion disease. Suspected cases are referred by clinicians and brain tissue undergoes immunohistochemistry and western blot analyses, all cases are therefore diagnosed with pathologically confirmed (definite) CJD. Genetic testing is performed through University Hospitals Department of Pathology.

### Control samples

See Supplementary Table 1 for sample sources and sizes. Control samples were chosen from genotype datasets available for biomedical research derived from countries that matched those providing case samples. In all but a single example, genotyping was done using Illumina arrays with substantial overlap with the Illumina Omniexpress array used to genotype cases.

### Quality control and imputation

Variant liftover to build GRCh37 was performed. Standard filters were applied using PLINK (v1.90b3v)^52^. Samples with a call rate below 98% (removed, n=116) and population outliers (established through population stratification with multidimensional scaling (MDS), removed=143) were rejected (Supplementary Figure 11). Cryptic duplicates and related samples (Pi_Hat > 0.1875 representing 3rd degree relatives) were excluded using IBS/IBD PLINK analysis with a 200K intersection marker set (removed, n=81). Individual SNPs were only kept if the following criteria were met: (a) genotype missing rate 1% in both cases and controls; (b) minor allele frequency >= 0.01; (c) consistency with Hardy-Weinberg disequilibrium by a degree-of-freedom goodness-of-fit test in controls (P>10^−4^); (d) not an A/T or G/C transversion; (e) no deviation from heterozygosity mean (±3 SD). Sex chromosomes were excluded. Phasing was performed using SHAPEIT^53^. Data were prepared for imputation by aligning the individual cohorts (in VCF format) to the 1000G Phase 3 Reference panel. The reference allele was designated as Allele1, and several tools were used to check correct alignment and compatibility with the Michigan Imputation Server pipeline (https://www.well.ox.ac.uk/~wrayner/tools/). Imputation was done using the Michigan Imputation Server with Minimac3 using the HRCr1.1 2016 reference panel assuming a mixed population^54^. All SNPs with an r^2^ threshold lower than 0.3 were excluded in the post-imputation QC analysis. The association test of 14 imputed cohorts was performed using the SNPTEST (v2.5.2) additive logistic regression model with 10 population covariates derived from MDS, retaining SNPs with (a) an “info” metric > 0.9 to remove poorly imputed SNPs, (b) a minor allele frequency above 0.005 and (c) a Hardy-Weinberg disequilibrium in controls. The final number of autosomal bi-allelic SNPs tested in the analysis was 6,314,477. For the replication phase the SNPs were genotyped using TaqMan assays, with sensitive allelic discrimination at all loci tested. P-values for the replication study were calculated from a fixed-effects meta-analysis of independent allele-based χ2 test for each case-control cohort (PLINK).

### Kuru and kuru resistance association testing

Meta analyses of kuru resistance was performed combining the results of two independent GWAS. 116 elderly ‘kuru-resistant’ women born before 1950 and aged >50 years, who did not develop kuru despite exposure, were compared to 31 individuals who died of kuru aged 25 or younger. This analysis included 122,622 variants after quality control procedures carried out in PLINK 1.9 (--maf 0.005 –hwe 1e-5 –geno 0.01). The second GWAS was performed comparing all other kuru exposed people aged >50 years, who did not develop kuru despite exposure (241 cases) to individuals born after kuru exposure, 1960 onwards (271 control individuals). This analysis had 123,672 (same quality control procedures as above) variants. Each association test was carried out using the –logistic function in PLINK 1.9 that performs a logistic regression based on an additive genetic model. Ten covariates were added to each analysis to help correct for population stratification in the regression analysis and obtained from PLINK 1.9 --pca function.

### Heritability estimation (SumHer)

We estimated SNP heritability using SumHer^9^, which uses GWAS summary statistics to estimate confounding bias and heritability providing greater flexibility by enabling the user to specify the heritability model, LDAK. The reference panel comprised 404 Finnish individuals from the 1000G reference panel. To account for systematic differences or distortion concerning the individuals in the study, we performed the analysis using prevalence (lifetime risk) of CJD 0.0002 and an ascertainment bias (case/control proportion) of 0.232.

### CAVIAR and eCAVIAR

CAVIAR utilises summary statistics to model association at a locus and estimates the posterior probability of a variant to be causal through modelling distribution conditional on the set of causal SNPs^14^. GWAS loci were defined as 100 SNPs upstream and downstream of the SNP with the highest association. LD matrix was calculated from sCJD GWAS data using PLINK --r2 function. CAVIAR was run with an undefined number of causal variants and causal set probability set at 95%. eCAVIAR is an extension of the CAVIAR framework to estimate the posterior probability of the same variant being causal in both GWAS and eQTL datasets^55^. The *STX6* locus was defined as previously and SNPs were pruned with variant inflation factor of 5 to extract distinct signals. We obtained eQTL data for 48 tissues included in the GTEx portal (v7). eQTLs for these 46 pruned variants with nominal P≤10^−5^ were defined as ‘eGenes’ and eCAVIAR performed for each in its target tissue. Resulting colocalisation posterior probability (CLPP) was ranked to identify target gene and tissue.

### Gene-based analysis (MAGMA / VEGAS2)

By aggregating the effects of all SNPs of a GWAS in a gene, correcting for LD and testing the joint association of all markers in the gene with the phenotype, it is possible to extend the power of detection and detect multiple weaker associations within a gene, which otherwise would be missed. Additionally, in gene-set analysis individual genes are aggregated to groups of genes sharing certain biological, functional or other characteristics and therefore pointing to involvement of specific biological pathways or cellular functions. MAGMA (version 1.06)^56^, based on a multiple regression model, and VEGAS2 (version 2v02)^57^, using simulations from the multivariate normal distribution, were employed for this analysis applying default parameters. The MAGMA model was the “snp-wise/mean” model, using the sum of –log(SNP p-value) as test statistic; for the VEGAS2 analysis we applied the Best-SNP test, which is most appropriate where only few SNPs regulate the gene of interest and the top SNP is in high LD with those SNPs. P values were adjusted using the Bonferroni method.

### PAINTOR

In addition, we utilised PAINTOR, a Bayesian probabilistic framework, which integrates GWAS summary and LD data with functional genomic annotation data such as ENCODE and FANTOM, to calculate posterior probabilities and identify a set of likely causal variants at any given risk locus (Supplementary Table 6). We aimed to identify causal variants 100kB around the lead SNP rs11586493 in the STX6 locus, our prime candidate identified in the GWAS, including all available functional brain tissue annotations (n=587).

### Whole exome sequencing

249 sCJD exomes were captured with SureSelect and HaloPlex (Agilent) kits. Exomes were sequenced on the Illumina HiSeq2000 platform with 100 bp paired-end runs. Sequences were aligned to the human reference genome using Novoalign software. The Genome Analysis Toolkit (GATK) Unified Genotyper were used for SNP and indel calling then sequences were recalibrated with the GATK Variant Recalibrator and variants annotated with ANNOVAR.

### Immunohistology

We used rabbit polyclonal anti-syntaxin-6 (Atlas Antibodies, HPA038558) and PDIA4 (Atlas Antibodies) to immunostain cases of sporadic CJD from archives of the Division of Neuropathology, University College London Hospitals NHS Foundation Trust. Immunostaining was performed on control cases and sCJD cases. Controls (non-neurodegenerative disease deaths, processed similarly to sCJD) were obtained from the Division of Neuropathology. A proportion of the samples originated from frozen material, and these were fixed by immersion into formalin, formic acid treated and then processed into a Formalin Fixed Paraffin Embedded (FFPE) sample. On all cases, frontal cortex and cerebellum (including the dentate nucleus) were stained.

### Measurement of reference and GWAS significant gene expression level in cerebellum by reverse transcription quantitative PCR (RT-qPCR)

RNA was extracted from 50-100 mg post-mortem cerebellum (10 sCJD cases and 10 neurologically normal controls) using the Zymo Direct-zol™ DNA/RNA Miniprep kit (Zymo Research, Irvine, US) according to the manufacturer’s instructions followed by DNAse I digestion and concentration using RNA Clean & Concentrator-5 columns (Zymo Research, Irvine, US) according to manufacturer’s instructions. All RNA samples were subsequently run on a TapeStation 2200 (Agilent, Santa Clara, US) using RNA ScreenTapes. RNA Integrity Number (RIN) was >4 for all samples tested. 1.6 µg total RNA from all samples was then reverse transcribed in 40 µl reactions using the Quantitect Reverse Transcription Kit (Qiagen, Hilden, Germany), followed by qPCR carried out using the Quantitect SYBR Green PCR Kit and Quantitect Primer Assays (both Qiagen, Hilden, Germany) according to the manufacturer’s instructions. Quantitect PCR Controls CYC1, RPL13 and UBE2D2 were used as reference genes, as recommended for post-mortem cerebellum by Rydbirk et al^23^, and all samples were run in triplicate on a QuantStudio 12K Flex (Life Technologies, Carlsbad, US) with dissociation curves to check for specific amplification. Qiagen primer assay reference numbers were as follows: CYC1 QT00209454; RPL13 QT00067963; UBE2D2 QT01009869; BMERB1/C16orf45 QT02319828; STX6 QT01679797; PDIA4 QT00015883; GAL3ST1 QT01013005; PRNP QT00070749.

### Generation of stable knockdown cells

N2aPK1/2 cells are highly susceptible to infection with Chandler/RML prions as previously described^25^. Cells were grown in Opti-MEM (Gibco), 10% FBS (Gibco), Penicillin-Streptomycin (100 U/ml and 100 µg/ml respectively; Gibco). The day prior to transfection, 5 × 10^5^ N2aPK1/2 cells were plated. Cells were transfected using Lipofectamine LTX with PLUS reagent (Invitrogen). 3.5 µg of pRetroSuper plasmid containing shRNA targeting mouse *Stx6* (5’-TGGAATGCTGGAGTGGCAGATCGCTATGG; Origene), *Prnp* (5’-GAGACAATCTAAACATTCT; as described previously^24^) or a non-effective scrambled 29mer (5’-GCACTACCAGAGCTAACTCAGATAGTACT; Origene). 24 h post-transfection cells were split 1:10 to integrate shRNA sequence. After 48 h, cells were placed under selection (3 µg/ml puromycin; Gibco). Clones were selected for high knockdown efficiency following *Stx6* shRNA transfection (‘shSTX6 1’, ‘shSTX6 2’) and scrambled shRNA (‘shScramble 1’, ‘shScramble 2’). *Prnp* knockdown cells were selected as a population due to high knockdown efficiency (‘shPRNP’). Gene expression measured after cells grown for 2 weeks in selection media.

### Scrapie cell assay

All cell lines were infected with Chandler/RML prions and cultured for up to 4x 1:8 splits before prion propagation was assayed as previously described^25^. Briefly, 1.8 × 10^4^ cells were seeded in a 96 well plate 24 h before infection with RML brain homogenate at 3 × 10^−6^ to 1 × 10^−8^ dilutions (12 wells per condition, n = 3). Cells were then split 1:8 every 3 to 4 days and assayed after the 3^rd^ and 4^th^ splits. Cells were fixed and lysed before proteinase K (PK) treatment and incubation with monoclonal anti-PrP antibody ICSM18 (D-Gen Ltd) followed by alkaline phosphatase-linked anti-IgG1 antiserum. Spots were visualized with alkaline phosphatase conjugate substrate (Bio-Rad) and PK-resistant PrP infected cells counted using the Bioreader 5000-Eβ (BioSys).

### Immunoblotting

Cells were lysed using RIPA buffer and protein quantified using Bradford assay (Sigma). Protein concentration was normalised in PBS. Lysates were mixed with 4 X SDS sample buffer and incubated at 95°C for 5 min. 65 µg of total protein was loaded onto a 4-12% Bis-Tris polyacrylamide gel (Invitrogen) and electrophoresed at 180 V for 1 h, then electroblotted to a nitrocellulose membrane (Invitrogen) at 35 V for 2 h. Membranes were blocked in Odyssey Blocking Buffer (LI-COR) for 1 h, probed with monoclonal anti-PrP antibody ICSM18 (1:500; D-Gen Ltd) or anti-Syntaxin 6 (1:500; Cell Signaling, 2869S) overnight and anti-Actin (1:5000, mouse monoclonal (Sigma; A5441) or 1:1000, rabbit polyclonal (Sigma; A2066)) for 1 h. After washing membranes were probed with fluorophore-conjugated secondary antibodies (LI-COR) for 1 h before imaging with Odyssey imaging system (LI-COR). Actin loading controls and all sample comparisons wecompre run on the same blot with molecular weight marker. Additional bands in lanes outside of those shown were removed for clarity.

## Data Availability

Summary statistics will be made available through the GWAS catalogue at NHGRI-EBI.

## Data availability

Summary statistics will be made available through the GWAS catalogue at NHGRI-EBI. Accession number not yet available.

## Acknowledgements

We thank Richard Newton for support with images. This study makes use of data generated by the Wellcome Trust Case-Control Consortium. A full list of the investigators who contributed to the generation of the data is available from www.wtccc.org.uk. Funding for the project was provided by the Wellcome Trust and Medical Research Council. We would like to thank patients, their families and carers, UK neurologists and other referring physicians, co-workers at the National Prion Clinic, our colleagues at the National Creutzfeldt-Jakob Disease Research and Surveillance Unit, Edinburgh. We thank the Director of the Papua New Guinea Institute of Medical Research, Professor Willie Pomat, and former Directors Professors Peter Siba and John Reeder, the staff of the PNGIMR, especially the kuru project field team, and the communities of the kuru-affected region for their generous support. We gratefully acknowledge the help of the late Carleton Gajdusek, the late Joseph Gibbs and their associates from the former Laboratory of Central Nervous System Studies of the National Institutes of Health, Bethesda, USA for archiving and sharing old kuru samples. The kuru studies were initially funded by a Wellcome Trust Principal Research Fellowship in the Clinical Sciences to JC, and since 2001, and all other aspects of the work by the Medical Research Council. Several authors at UCL/UCLH receive funding from the Department of Health’s NIHR Biomedical Research Centres funding scheme. Some of this work was supported by the Department of Health funded National Prion Monitoring Cohort study. SJC receives an NHMRC Practitioner Fellowship (ID# APP1105784). Tze How Mok is supported by a Fellowship award from Alzheimer’s Society, UK (grant number 341 (AS-CTF-16b-007)) and CJD Support Network UK Research Support Grants. Funding for the collection of Polish samples for study was provided Medical University of Lodz. The Italian national surveillance of Creutzfeldt-Jakob disease and related disorders is partially supported by the Ministero della Salute, Italy. The German National Reference Centre for TSE is funded by grants from the Robert-Koch-Institute. The Dutch National Prion Disease Registry is funded by the National Institute for Public Health and the Environment (RIVM), which is part from the Ministry for Health, Welfare and Sports, The Netherlands. PS-J was supported by Instituto de Salud Carlos III [Fondo de Investigación Sanitaria, PI16/01652] Accion Estrategica en Salud integrated in the Spanish National I+D+i Plan and financed by Instituto de Salud Carlos III (ISCIII)– Subdireccion General de Evaluacion and the Fondo Europeo de Desarrollo Regional (FEDER – “Una Manera de Hacer Europa”). We thank Inés Santiuste and the Valdecilla Biobank (PT17/0015/0019), integrated in the Spanish Biobank Network, for their support and collaboration in sample collection and management. The study on Italian controls was supported by the Ministero dell’Istruzione, dell’Università e della Ricerca – MIUR project “Dipartimenti di Eccellenza 2018 – 2022” (n° D15D18000410001) to the Department of Medical Sciences, University of Torino (G.M.) and the AIRC – Associazione Italiana per la Ricerca sul Cancro (IG 2018 Id.21390 to G.M.). The Three-City Study was performed as part of a collaboration between the Institut National de la Santé et de la Recherche Médicale (Inserm), the Victor Segalen Bordeaux II University and Sanofi-Synthélabo. The Fondation pour la Recherche Médicale funded the preparation and initiation of the study. The 3C Study was also funded by the Caisse Nationale Maladie des Travailleurs Salariés, Direction Générale de la Santé, MGEN, Institut de la Longévité, Agence Française de Sécurité Sanitaire des Produits de Santé, the Aquitaine and Bourgogne Regional Councils, Agence Nationale de la Recherche, ANR supported the COGINUT and COVADIS projects. Fondation de France and the joint French Ministry of Research/INSERM «Cohortes et collections de données biologiques» programme. Lille Génopôle received an unconditional grant from Eisai. The Three-city biological bank was developed and maintained by the laboratory for genomic analysis LAG-BRC - Institut Pasteur de Lille. This work was also funded by the Pasteur Institut de Lille, the Lille Métropole Communauté Urbaine, the Haut-de France council, the European Community (FEDER) and the French government’s LABEX DISTALZ program (development of innovative strategies for a transdisciplinary approach to Alzheimer’s disease). The French National Surveillance Network for Creutzfeldt-Jakob disease is supported by Santé Publique France.

## Notes

### Competing Interest Statement

The authors have declared no competing interest.

## References

1 Collinge, J. Prion diseases of humans and animals: their causes and molecular basis. Annual Review of Neuroscience 24, 519–550 (2001).

2 Collinge, J. Variant Creutzfeldt-Jakob disease. Lancet 354, 317–323, doi:10.1016/S0140-6736(99)05128-4 (1999).

3 Mead, S., Lloyd, S. & Collinge, J. Genetic Factors in Mammalian Prion Diseases. Annu Rev Genet 53, 117–147, doi:10.1146/annurev-genet-120213-092352 (2019).

4 NCJDRSU. Annual report. (2017).

5 Webb, T. E. et al. First Report of Creutzfeldt-Jakob Disease Occurring in 2 Siblings unexplained by PRNP mutation. J Neuropathol Exp Neurol 67, 838–841 (2008).

6 Mead, S. et al. Genome-wide association study in multiple human prion diseases suggests genetic risk factors additional to PRNP. Hum Mol Genet 21, 1897–1906 (2011).

7 Sanchez-Juan, P. et al. A genome wide association study links glutamate receptor pathway to sporadic Creutzfeldt-Jakob disease risk. PLoS One 10, e0123654, doi:10.1371/journal.pone.0123654 (2014).

8 Mead, S. et al. Genetic risk factors for variant Creutzfeldt-Jakob disease: a genome-wide association study. Lancet Neurol 8, 57–66 (2009).

9 Speed, D. & Balding, D. J. SumHer better estimates the SNP heritability of complex traits from summary statistics. Nat Genet 51, 277–284, doi:10.1038/s41588-018-0279-5 (2019).

10 Yang, J., Lee, S. H., Goddard, M. E. & Visscher, P. M. GCTA: a tool for genome-wide complex trait analysis. Am J Hum Genet 88, 76–82, doi:10.1016/j.ajhg.2010.11.011 (2011).

11 Collinge, J. & Clarke, A. R. A general model of prion strains and their pathogenicity. Science 318, 930–936, doi:10.1126/science.1138718 (2007).

12 Mead, S. et al. Clinical Trial Simulations Based on Genetic Stratification and the Natural History of a Functional Outcome Measure in Creutzfeldt-Jakob Disease. JAMA Neurol (2016).

13 Thompson, A. G. et al. The Medical Research Council Prion Disease Rating Scale: a new outcome measure for prion disease therapeutic trials developed and validated using systematic observational studies. Brain 136, 1116–1127 (2013).

14 Hormozdiari, F., Kostem, E., Kang, E. Y., Pasaniuc, B. & Eskin, E. Identifying causal variants at loci with multiple signals of association. Genetics 198, 497–508, doi:10.1534/genetics.114.167908 (2014).

15 Mead, S. et al. Sporadic--but not variant--Creutzfeldt-Jakob disease is associated with polymorphisms upstream of PRNP exon 1. Am J Hum Genet 69, 1225–1235, doi:10.1086/324710 (2001).

16 Bratosiewicz-Wasik, J. et al. Association between the PRNP 1368 polymorphism and the occurrence of sporadic Creutzfeldt-Jakob disease. Prion 6, 413–416, doi:10.4161/pri.21773 (2012).

17 Sanchez-Juan, P. et al. A polymorphism in the regulatory region of PRNP is associated with increased risk of sporadic Creutzfeldt-Jakob disease. BMC Med Genet 12, 73, doi:10.1186/1471-2350-12-73 (2011).

18 Vollmert, C. et al. Significant association of a M129V independent polymorphism in the 5’ UTR of the PRNP gene with sporadic Creutzfeldt-Jakob disease in a large German case-control study. J Med Genet 43, e53, doi:10.1136/jmg.2006.040931 (2006).

19 Consortium, G. T. The Genotype-Tissue Expression (GTEx) project. Nat Genet 45, 580–585, doi:10.1038/ng.2653 (2013).

20 Meissner, B. et al. MRI and clinical syndrome in dura materrelated Creutzfeldt-Jakob disease. J Neurol (2009).

21 Kichaev, G. et al. Integrating functional data to prioritize causal variants in statistical finemapping studies. PLoS Genet 10, e1004722, doi:10.1371/journal.pgen.1004722 (2014).

22 Koriath, C. et al. Predictors for a dementia gene mutation based on gene-panel next-generation sequencing of a large dementia referral series. Mol Psychiatry, doi:10.1038/s41380-018-0224-0 (2018).

23 Rydbirk, R. et al. Assessment of brain reference genes for RT-qPCR studies in neurodegenerative diseases. Sci Rep 6, 37116, doi:10.1038/srep37116 (2016).

24 Brown, C. A. et al. In vitro screen of prion disease susceptibility genes using the scrapie cell assay. Hum Mol Genet 23, 5102–5108, doi:10.1093/hmg/ddu233 (2014).

25 Klohn, P. C., Stoltze, L., Flechsig, E., Enari, M. & Weissmann, C. A quantitative, highly sensitive cell-based infectivity assay for mouse scrapie prions. Proc Natl Acad Sci U S A 100, 11666–11671, doi:10.1073/pnas.1834432100 (2003).

26 Goold, R. et al. Rapid cell-surface prion protein conversion revealed using a novel cell system. Nat Commun 2, 281 (2011).

27 Claussnitzer, M. et al. A brief history of human disease genetics. Nature 577, 179–189, doi:10.1038/s41586-019-1879-7 (2020).

28 Wendler, F. & Tooze, S. Syntaxin 6: the promiscuous behaviour of a SNARE protein. Traffic 2, 606–611, doi:10.1034/j.1600-0854.2001.20903.x (2001).

29 Yamasaki, T., Suzuki, A., Hasebe, R. & Horiuchi, M. Retrograde Transport by Clathrin-Coated Vesicles is Involved in Intracellular Transport of PrP(Sc) in Persistently Prion-Infected Cells. Sci Rep 8, 12241, doi:10.1038/s41598-018-30775-1 (2018).

30 Yim, Y. I. et al. The multivesicular body is the major internal site of prion conversion. J Cell Sci (2015).

31 Bartoletti-Stella, A. et al. Analysis of RNA Expression Profiles Identifies Dysregulated Vesicle Trafficking Pathways in Creutzfeldt-Jakob Disease. Mol Neurobiol 56, 5009–5024, doi:10.1007/s12035-018-1421-1 (2019).

32 Goold, R., McKinnon, C. & Tabrizi, S. J. Prion degradation pathways: Potential for therapeutic intervention. Mol Cell Neurosci (2015).

33 Collinge, J. Mammalian prions and their wider relevance in neurodegenerative diseases. Nature 539, 217–226, doi:10.1038/nature20415 (2016).

34 Colin, M. et al. From the prion-like propagation hypothesis to therapeutic strategies of anti-tau immunotherapy. Acta Neuropathol 139, 3–25, doi:10.1007/s00401-019-02087-9 (2020).

35 Chen, J. A. et al. Joint genome-wide association study of progressive supranuclear palsy identifies novel susceptibility loci and genetic correlation to neurodegenerative diseases. Mol Neurodegener 13, 41, doi:10.1186/s13024-018-0270-8 (2018).

36 Hoglinger, G. U. et al. Identification of common variants influencing risk of the tauopathy progressive supranuclear palsy. Nat Genet 43, 699–705, doi:10.1038/ng.859 (2011).

37 Takahashi, T. & Suzuki, T. Role of sulfatide in normal and pathological cells and tissues. J Lipid Res 53, 1437–1450, doi:10.1194/jlr.R026682 (2012).

38 Platt, F. M., d’Azzo, A., Davidson, B. L., Neufeld, E. F. & Tifft, C. J. Lysosomal storage diseases. Nat Rev Dis Primers 4, 27, doi:10.1038/s41572-018-0025-4 (2018).

39 Honke, K. et al. Paranodal junction formation and spermatogenesis require sulfoglycolipids. Proc Natl Acad Sci U S A 99, 4227–4232, doi:10.1073/pnas.032068299 (2002).

40 Klein, T. R., Kirsch, D., Kaufmann, R. & Riesner, D. Prion rods contain small amounts of two host sphingolipids and revealed by thin-layer chromatography and mass spectrometry. Biol Chem 379, 666 (1998).

41 Agostini, F. et al. Prion protein accumulation in lipid rafts of mouse aging brain. PLoS ONE 8, e74244 (2013).

42 Hwang, D. et al. A systems approach to prion disease. Mol Syst Biol 5, 252 (2009).

43 Bremer, J. et al. Axonal prion protein is required for peripheral myelin maintenance. Nat Neurosci (2010).

44 Kuffer, A. et al. The prion protein is an agonistic ligand of the G protein-coupled receptor Adgrg6. Nature 536, 464–468 (2016).

45 Skedsmo, F. S. et al. Demyelinating polyneuropathy in goats lacking prion protein. FASEB J, doi:10.1096/fj.201902588R (2019).

46 Wang, S. B. et al. Protein Disulfide Isomerase Regulates Endoplasmic Reticulum Stress and the Apoptotic Process during Prion Infection and PrP Mutant-Induced Cytotoxicity. PLoS ONE 7, e38221 (2012).

47 Rai, A. et al. bMERB domains are bivalent Rab8 family effectors evolved by gene duplication. Elife 5, doi:10.7554/eLife.18675 (2016).

48 Zerr, I. et al. Updated clinical diagnostic criteria for sporadic Creutzfeldt-Jakob disease. Brain 132, 2659–2668 (2009).

49 Collins, S. et al. Creutzfeldt-Jakob disease in Australia 1970-1999. Neurology 59, 1365–1371, doi:10.1212/01.wnl.0000031793.11602.8c (2002).

50 Collins, S. J. et al. No evidence for prion protein gene locus multiplication in Creutzfeldt-Jakob disease. Neurosci Lett 472, 16–18, doi:10.1016/j.neulet.2010.01.043 (2010).

51 Mead, S. et al. Genome-wide association study in multiple human prion diseases suggests genetic risk factors additional to PRNP. Hum Mol Genet 21, 1897–1906 (2012).

52 Purcell, S. et al. PLINK: A tool set for whole-genome association and population-based linkage analyses. American Journal of Human Genetics 81, 559–575 (2007).

53 Delaneau, O., Marchini, J. & Zagury, J. F. A linear complexity phasing method for thousands of genomes. Nat Methods 9, 179–181, doi:10.1038/nmeth.1785 (2011).

54 Das, S. et al. Next-generation genotype imputation service and methods. Nat Genet 48, 1284–1287, doi:10.1038/ng.3656 (2016).

55 Hormozdiari, F. et al. Colocalization of GWAS and eQTL Signals Detects Target Genes. Am J Hum Genet 99, 1245–1260, doi:10.1016/j.ajhg.2016.10.003 (2016).

56 de Leeuw, C. A., Mooij, J. M., Heskes, T. & Posthuma, D. MAGMA: generalized gene-set analysis of GWAS data. PLoS Comput Biol 11, e1004219, doi:10.1371/journal.pcbi.1004219 (2015).

57 Mishra, A. & Macgregor, S. VEGAS2: Software for More Flexible Gene-Based Testing. Twin Res Hum Genet 18, 86–91, doi:10.1017/thg.2014.79 (2015).

